# Deep Learning-Based Recognizing COVID-19 and other Common Infectious Diseases of the Lung by Chest CT Scan Images

**DOI:** 10.1101/2020.03.28.20046045

**Authors:** Min Fu, Shuang-Lian Yi, Yuanfeng Zeng, Feng Ye, Yuxuan Li, Xuan Dong, Yan-Dan Ren, Linkai Luo, Jin-Shui Pan, Qi Zhang

**Author notes:** Correspondence: Qi Zhang, MD, PhD., Department of Nosocomial Infection and Public Health, Jin Yin-Tan Hospital, Wuhan 430023, No. 1, Yintan Road, Jin-Shui Pan, MD, PhD., Department of Gastroenterology, Zhongshan Hospital Affiliated to Xiamen University No. 201-209, Hubin Nan Road, Xiamen 361004, Fujian, China, Linkai Luo, Prof., School of Aerospace Engineering, Xiamen University, Xiang’an Nan Road, Xiamen 361102, Fujian, China. The authors contribute equally to the work (Min Fu, Shuang-Lian Yi, Yuanfeng Zeng, Feng Ye). **E-mails:** Min Fu.

## Abstract

**Purpose:** COVID-19 has become global threaten. CT acts as an important method of diagnosis. However, human–based interpretation of CT imaging is time consuming. More than that, substantial inter-observer-variation cannot be ignored. We aim at developing a diagnostic tool for artificial intelligence (AI)–based classification of CT images for recognizing COVID-19 and other common infectious diseases of the lung.

**Experimental Design:** In this study, images were retrospectively collected and prospectively analyzed using machine learning. CT scan images of the lung that show or do not show COVID-19 were used to train and validate a classification framework based on convolutional neural network. Five conditions including COVID-19 pneumonia, non-COVID-19 viral pneumonia, bacterial pneumonia, pulmonary tuberculosis, and normal lung were evaluated. Training and validation set of images were collected from Wuhan Jin Yin-Tan Hospital whereas test set of images were collected from Zhongshan Hospital Xiamen University and the fifth Hospital of Wuhan.

**Results:** Accuracy, sensitivity, and specificity of the AI framework were reported. For test dataset, accuracies for recognizing normal lung, COVID-19 pneumonia, non-COVID-19 viral pneumonia, bacterial pneumonia, and pulmonary tuberculosis were 99.4%, 98.8%, 98.5%, 98.3%, and 98.6%, respectively. For the test dataset, accuracy, sensitivity, specificity, PPV, and NPV of recognizing COVID-19 were 98.8%, 98.2%, 98.9%, 94.5%, and 99.7%, respectively.

**Conclusions:** The performance of the proposed AI framework has excellent performance of recognizing COVID-19 and other common infectious diseases of the lung, which also has balanced sensitivity and specificity.

## Introduction

Coronaviruses are non-segmented positive-sense RNA viruses with envelope that belongings to the family Coronaviridae, which widely distributed in humans and other mammals. Since the beginning of this century, coronavirus has caused several localized epidemics and even global pandemics, such as SARS, Middle East Respiratory Syndrome, and the ongoing coronavirus disease 2019 (COVID-19). Up to 27 March 2020, 509164 confirmed cases were reported with 23335 deaths worldwide; over 82078 cases of COVID-19 have been confirmed in mainland China, with a mortality rate of 4.0%^(1)^. Although the upward trend of COVID-19 has been effectively curbed, the number of confirmed cases has increased dramatically in several countries, such as South Korea, Japan, Italy, and other countries. According to the WHO interim guidance^(2)^ and series diagnosis and treatment scheme for COVID-19 of China, confirmed case of COVID-19 was made on the basis of a positive result on high-throughput sequencing or real-time reverse-transcriptase–polymerase-chain-reaction (RT-PCR) assay of specimen collected by nasal and pharyngeal swab. However, the false negative rate of nucleic acid detection may be relatively high in early stage of COVID-19. Sometimes, repeated tests are needed to get positive results. Novel coronavirus that leads to COVID-19 is encoded by RNA, which is a highly unstable and tends to be degraded by RNAase. RNAase is widely found in saliva and surrounding environment. Thus, RNA of novel coronavirus in the specimen collected by nasal and pharyngeal swab may have been degraded by contaminated RNAase, which at least partly explains the low positive rate of nucleic acid assay from nasal and pharyngeal swab. Another constraint in practice is that the supply of assay kits of nucleic acid detection may be seriously inadequate in case of a large-scale outbreak of disease. In contrast, COVID-19 has relatively unique imaging features in CT manifestations. In early stage (less than 1 week after symptom onset), the predominant pattern was unilateral or bilateral ground-glass opacities. Within 1–3 weeks, ground-glass opacities will progress to or co-existed with consolidations.^(3)^ According to the investigation by Guan *et al*,^(4)^ at the time of admission, 86.2% revealed abnormal CT scans whereas radiographic or CT abnormality was found in 97.1% of the patients with severe type of COVID-19. What cause the characteristic abnormality found by CT scans? Histological examination reveals diffuse alveolar damage with cellular fibromyxoid exudates, which may lead to the changes in CT scans^(5)^. Fibromyxoid exudates in alveoli may further cause disorder in gas exchange and even respiratory failure, which is consistent with the observation by Li *et al*.^(6)^

Given the rapid spread of COVID-19 and the above advantages of CT scan, we developed deep learning-based detection of characteristic abnormality to facilitate the early diagnosis of COVID-19.

## Methods

### Patients

Spiral CT scanning of the lung was performed in Department of radiology, Wuhan Jin Yin-Tan Hospital, Zhongshan Hospital Xiamen University, the fifth Hospital of Wuhan between January 1, 2015 and February 29, 2020. Adult patients who aged between 18 and 75 were enrolled in case of the following conditions: laboratory confirmed COVID-19, non-COVID-19 viral pneumonia, bacterial pneumonia, pulmonary tuberculosis, or absent from abnormal finding in lung CT (normal lung). Case of COVID-19 was confirmed based on the positive result of fluorescent RT-PCR analysis of COVID-19 nucleic acid detection. Apart from COVID-19, the diagnoses of other diseases were made based on pathogen examinations according to the relevant guidelines, and were further confirmed by clinical manifestations, and treatment outcomes. For the patients with more than one kind of the fore-mentioned diseases, such as bacterial pneumonia complicated with pulmonary tuberculosis, will be ruled out from this study. Findings of CT scan images, results of pathogen examinations, and clinical diagnoses were recorded. This study was approved by the Ethics Commission of Zhongshan Hospital Xiamen University. Written informed consent was waived by the Ethics Commission of the designated hospital because of non-interventional study and no identifiable personal information was recorded.

### Images

Lung CT scan images from the enrolled patients were retrospectively collected. Images without perfect lung fields were filtered out. Identifiable personal information, such as name of the enrolled patients, name of hospital, etc, was removed. Consecutive images of the lung fields for each patient were selected for image recognizing. For a specific patient, he will be classified as “COVID-19” case if typical CT manifestations related to COVID-19 were identified even in only one image. This rule was also true for other diseases, such as non-COVID-19 virtual pneumonia, biological pneumonia, and pulmonary tuberculosis. But for each selectee, only when all of his images were recognized as “normal” will he be classified into the group of “normal”.

### Datasets

Lung CT scan images collected from Department of radiology, Wuhan Jin Yin-Tan Hospital were randomly divided into training set or validation set at a ratio of 3:1. The training set was employed to construct the AI model whereas the validation set was used to assess the accuracy of classification performance of the constructed model. This process was repeated for 5 times. Lung CT scan images collected from Department of radiology, Zhongshan Hospital Xiamen University, the fifth Hospital of Wuhan acted as test sets to evaluate the generalization performance in classifying the images beyond the lung CT scan images used in the training set or validation set.

### Training and validation the algorithm

Based on deep learning, we used the PyTorch platform to adopt the ResNet-50 architecture pretrained using the ImageNet dataset^(7)^ to develop our AI algorithm. The retraining consisted of initializing the convolutional layers with loaded pretrained weights and updating of the neural network to recognize our classes such as COVID-19, non-COVID-19 viral pneumonia, bacterial pneumonia, pulmonary tuberculosis, or and normal lung. The network structure was kept unchanged in this study. However, the weights of the last fully connected layer and the last three convolutional layers were tuned. Firstly, the weights were updated by Adam optimizer and the learning rate was 0.0001; Secondly, the weights were updated by SGD optimizer while the learning rate was set as 0.001. This strategy was superior to a sole optimizer such as SGD or Adam optimizer. After 50 epochs (iterations through the entire dataset), the training was stopped if no further improvement in accuracy or cross-entropy loss were observed. Schematic diagram for the development of the AI algorithm was shown in Supplementary Figure 1.

**Supplementary Figure 1.**
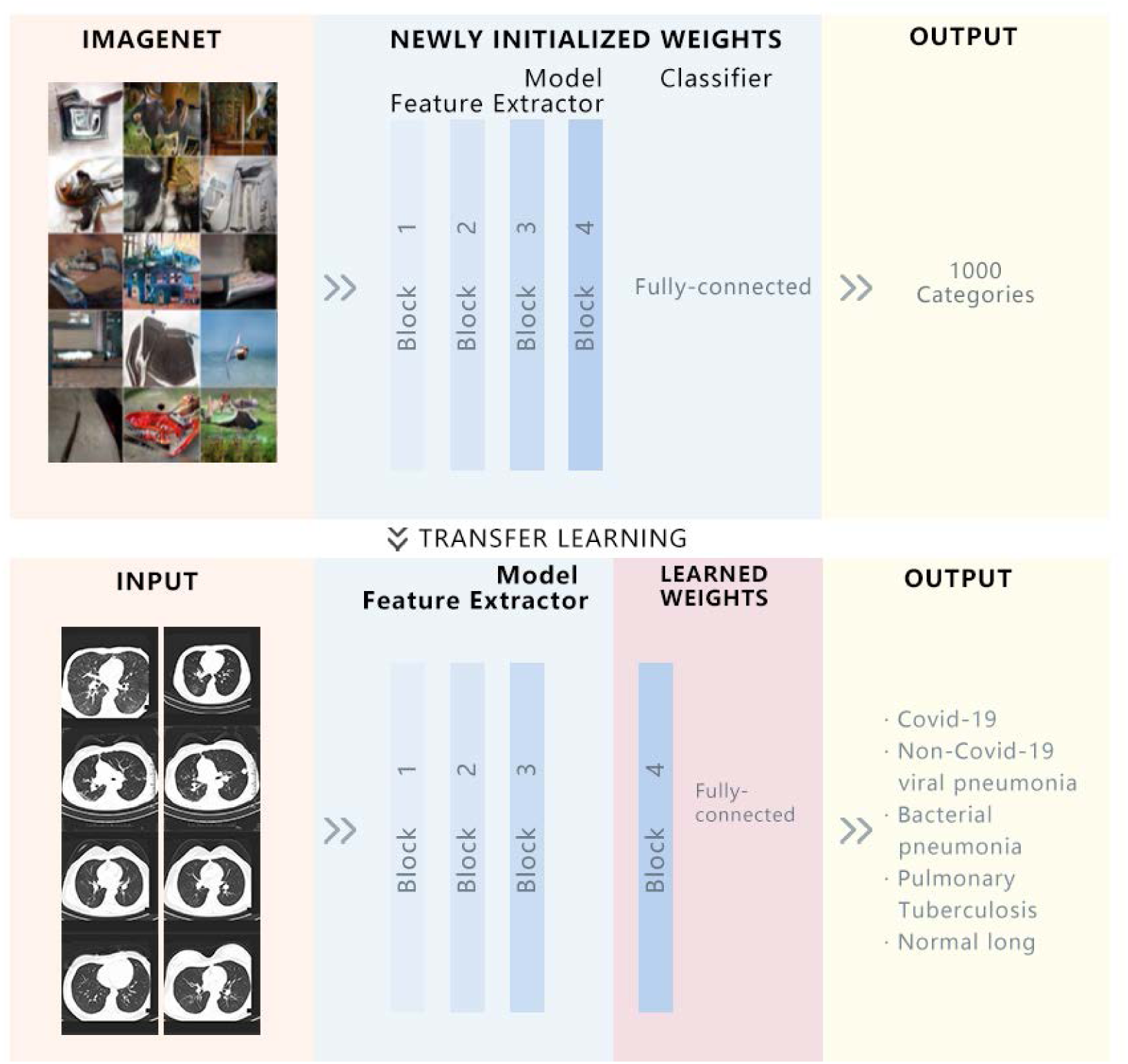
Schematic diagram for the development of the AI algorithm.

### Testing of the AI algorithm

The AI algorithm was further tested by the lung CT scan images collected from Department of radiology, Zhongshan Hospital Xiamen University, and the Fifth Hospital of Wuhan. The classification performance was evaluated independently in the images collected from these two hospitals.

### Comparison between the AI algorithm and radiologist

The lung CT scan images collected from Department of radiology, Zhongshan Hospital Xiamen University, and the Fifth Hospital of Wuhan were also sent to expert radiologist to make a diagnosis. Classification performance and cost of time were compared with that of the AI algorithm. Expert radiologists were senior staffs of Department of Radiology, Zhongshan Hospital Xiamen University, with clinical experience about 10 years. Diagnosis was made independently.

### Statistical analysis

To evaluate the classification performance of the AI algorithm on lung CT scan images, five indies including AUC, accuracy, sensitivity, specificity, PPV, and NPV were calculated. The receiver operating characteristics (ROC) curves plot the true positive rate (sensitivity) versus the false positive rate (1-specificity). P < 0.05 was set as the level for statistical significance for two-tailed paired test.

## Results

### Characteristics of patient and image

After filtering those images without good lung fields. Three radiologists with more than 10 years of clinical experience labelled infection lesions in the images, and lesions reach a consensus were labelled. A total of 60427 CT scan images collected from Wuhan Jin Yin-Tan Hospital from the following patients: 100 cases of COVID-19 pneumonia, 102 cases of non-COVID-19 viral pneumonia, 103 cases of bacterial pneumonia, 105 cases of pulmonary tuberculosis, 200 cases of normal lung, were employed to develop the model (Table 1). These images were randomly divided into training and validation datasets. Enrolled images in training and validation dataset covered almost all common types of infectious diseases of the lung.

**Table 1.** Characteristics of the enrolled patients and images.

|  | Training and Validation set |  | Test set |  | Test set |  |
| --- | --- | --- | --- | --- | --- | --- |
|  | Wuhan Jin Yin-Tan Hospital |  | Zhongshan Hospital Xiamen University |  | the fifth Hospital of Wuhan |  |
|  | Patients | Images | Patients | Images | Patients | Images |
| Normal | 200 | 19976 | 50 | 4478 | 50 | 4419 |
| **COVID-19** | 100 | 10057 | 13 | 1288 | 37 | 3599 |
| **Non-COVID-19**<br>**viral pneumonia** | 102 | 10028 | 32 | 3004 | 20 | 2101 |
| **Bacterial**<br>**pneumonia** | 103 | 10107 | 28 | 2719 | 25 | 2386 |
| **Pulmonary**<br>**tuberculosis** | 105 | 10259 | 16 | 1589 | 38 | 3618 |

**Supplementary Table 1.** Diagnostic performance of the AI algorithm during validation.

|  | Accuracy (%) | Sensitivity (%) | Specificity (%) | PPV (%) | NPV (%) |
| --- | --- | --- | --- | --- | --- |
| **Normal** | 99.2 | 99.0 | 99.3 | 98.5 | 99.5 |
| **COVID-19** | 98.9 | 96.7 | 99.3 | 96.7 | 99.3 |
| **Non-COVID-19**<br>**viral pneumonia** | 98.9 | 96.1 | 99.4 | 97.0 | 99.2 |
| **Bacterial**<br>**pneumonia** | 98.6 | 96.0 | 99.2 | 95.8 | 99.2 |
| **Pulmonary**<br>**tuberculosis** | 99.1 | 97.2 | 99.4 | 97.3 | 99.4 |

Note: PPV, positive predictive value; NPV, negative predictive value.

### Performance of the AI algorithm during training and validation

Based on validation dataset, we evaluated the performance of our AI algorithm in diagnosing the most common infectious diseases of the lung, including non-COVID-19 viral pneumonia, bacterial pneumonia, pulmonary tuberculosis except COVID-19. During training and validation process, accuracy and cross-entropy were plotted against the iteration step, which were shown in Figure 1. Confusion matrix of the AI framework during validation process was also shown in Figure 1. Multi-class comparison was performed between COVID-19, non-COVID-19 viral pneumonia, bacterial pneumonia, pulmonary tuberculosis, and normal lung. Binary comparison between COVID-19 and the other four types, including non-COVID-19 viral pneumonia, bacterial pneumonia, pulmonary tuberculosis and normal lung, was also implemented to evaluate the performance of recognizing COVID-19. Accuracy, sensitivity, specificity, PPV, and NPV of recognizing COVID-19 were 98.9%, 96.7%, 99.3%, 96.7%, and 99.3%, respectively (Supplementary Table 1). Similarly, binary comparison was performed for the other four conditions (Supplementary Table 1).

**Figure 1.**
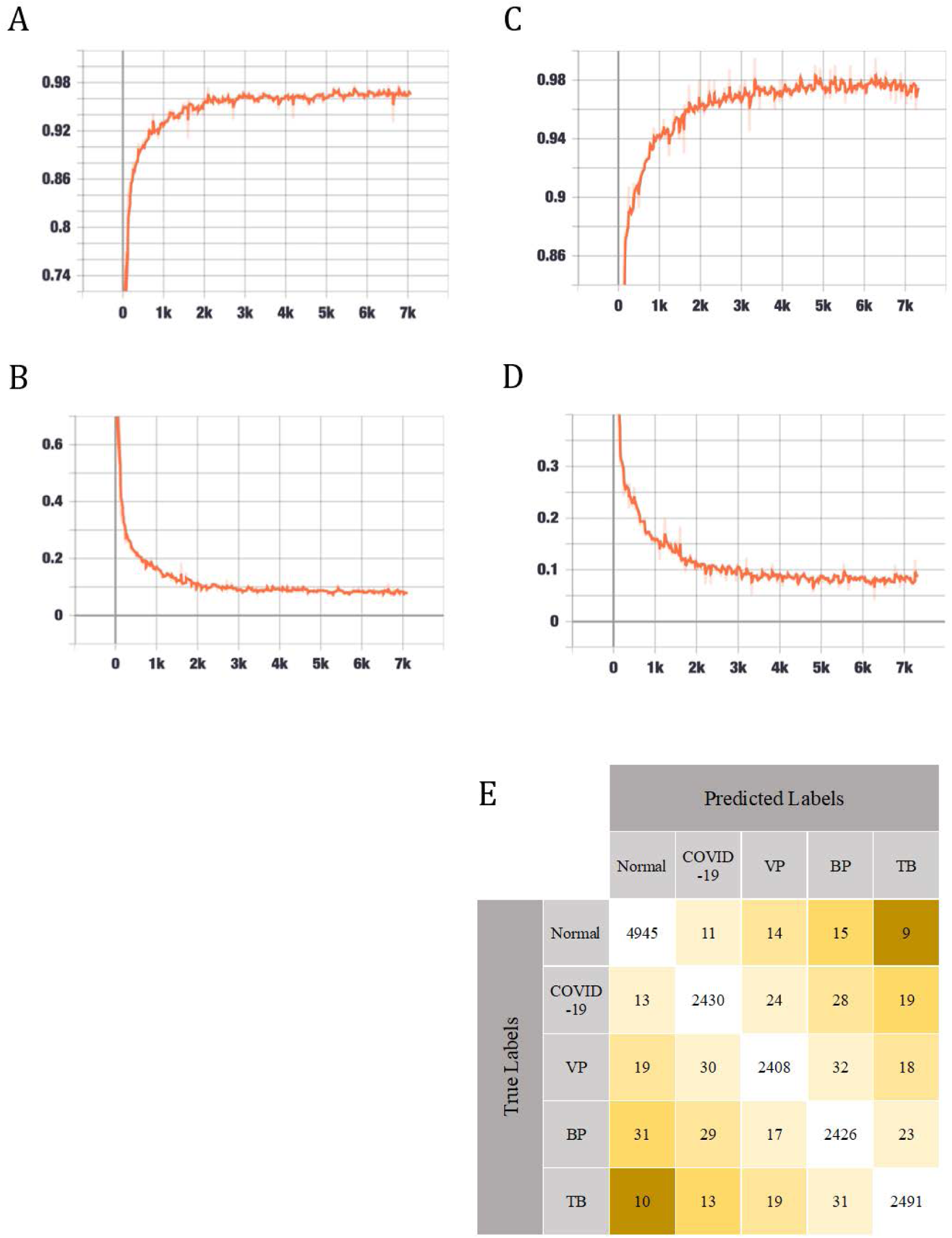
Performance of the AI algorithm during training and validation. (A) Classification accuracy is plotted against training epochs. (B) The categorical cross-entropy loss is shown as a function of training epochs for the binary classification problem. (C) Classification accuracy is plotted against validation epochs. (D) The categorical cross-entropy loss is shown as a function of validation epochs for the binary classification problem. The curve is smoothed. (E) Confusion matrix of the AI framework during validation process.

### Performance of the AI algorithm during test

From Zhongshan Hospital Xiamen University, and the fifth Hospital of Wuhan, 29201 CT scan images were collected from the following patients: 50 cases of COVID-19 pneumonia, 52 cases of non-COVID-19 viral pneumonia, 53 cases of bacterial pneumonia, 54 cases of pulmonary tuberculosis, 100 cases of normal lung, were employed to develop the model (Table 1). Multi-class comparison was performed between COVID-19, non-COVID-19 viral pneumonia, bacterial pneumonia, pulmonary tuberculosis, and normal lung. Confusion matrix of the AI framework based on test dataset was shown in Figure 2. Binary classification between COVID-19 and the other four types, including non-COVID-19 viral pneumonia, bacterial pneumonia, pulmonary tuberculosis and normal lung, was also implemented to evaluate the performance of recognizing COVID-19. For test dataset, accuracy, sensitivity, specificity, PPV, and NPV of recognizing COVID-19 were 98.8%, 98.2%, 98.9%, 94.5%, and 99.7%, respectively (Supplementary Table 2). The ROC curve was generated to evaluate the AI algorithm’s ability to distinguish COVID-19 from other four types. The area under the ROC curve was 99.0% (Figure 2).

**Supplementary Table 2.**
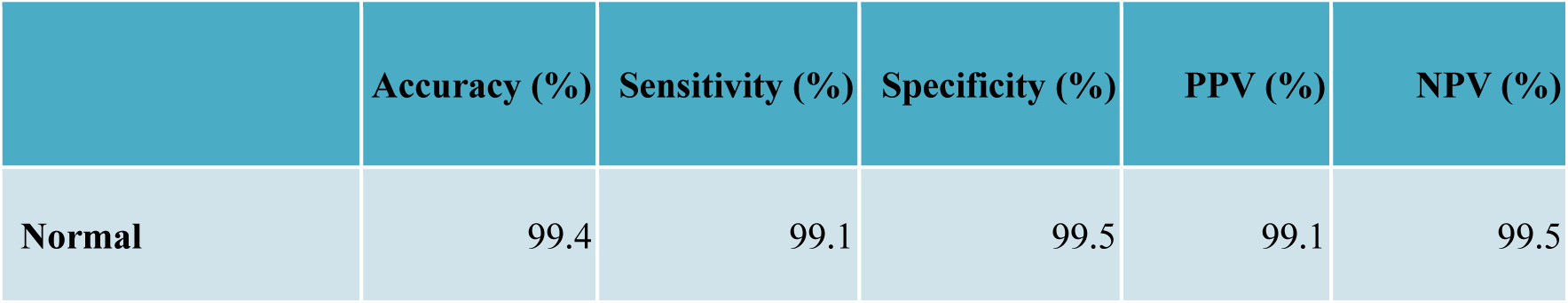
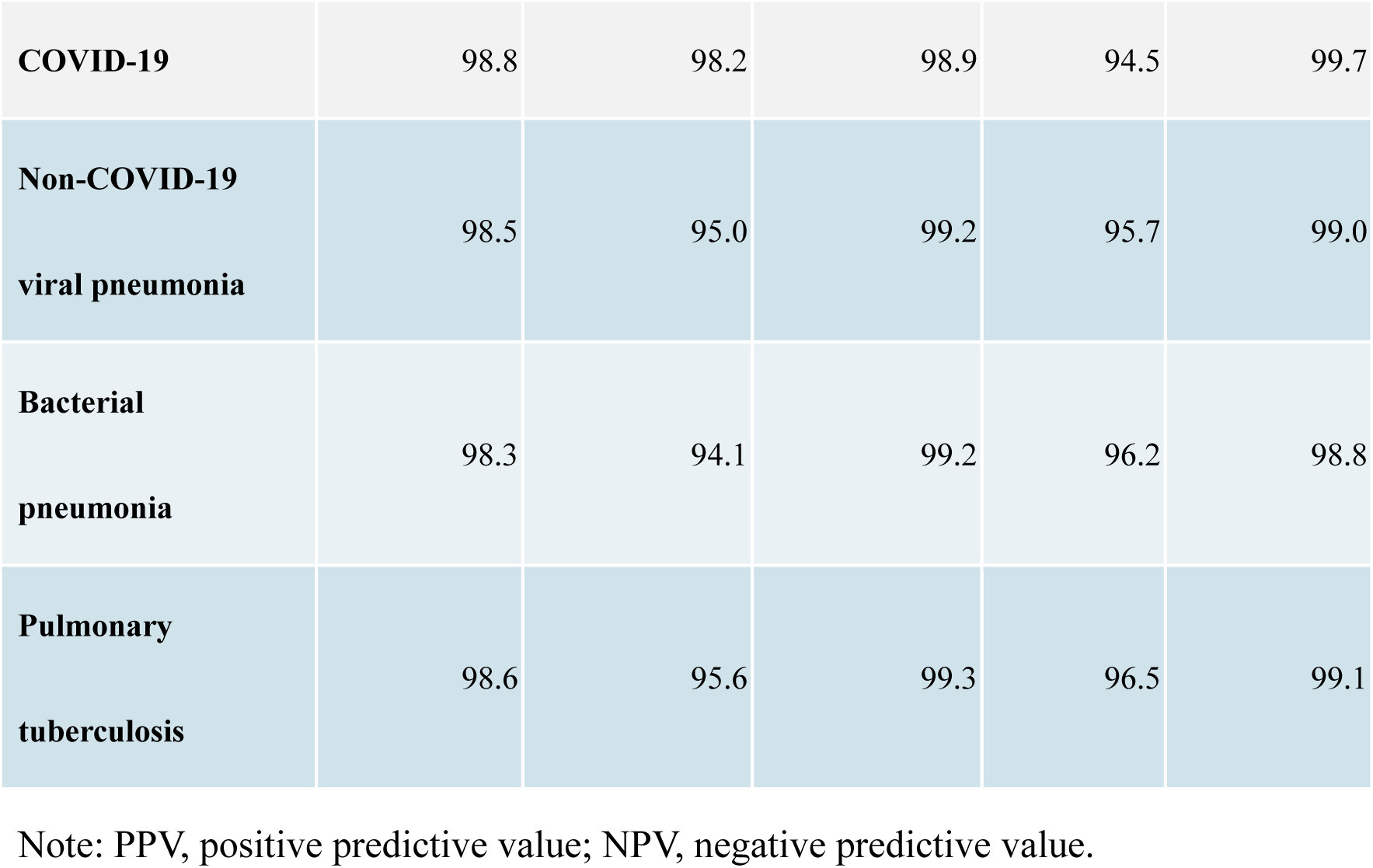
Diagnostic performance of the AI algorithm during test.

|  | Accuracy (%) | Sensitivity (%) | Specificity (%) | PPV (%) | NPV (%) |
| --- | --- | --- | --- | --- | --- |
| Normal | 99.4 | 99.1 | 99.5 | 99.1 | 99.5 |
| **COVID-19** | 98.8 | 98.2 | 98.9 | 94.5 | 99.7 |
| **Non-COVID-19**<br>**viral pneumonia** | 98.5 | 95.0 | 99.2 | 95.7 | 99.0 |
| **Bacterial**<br>**pneumonia** | 98.3 | 94.1 | 99.2 | 96.2 | 98.8 |
| **Pulmonary**<br>**tuberculosis** | 98.6 | 95.6 | 99.3 | 96.5 | 99.1 |

Note: PPV, positive predictive value; NPV, negative predictive value.

**Figure 2.**
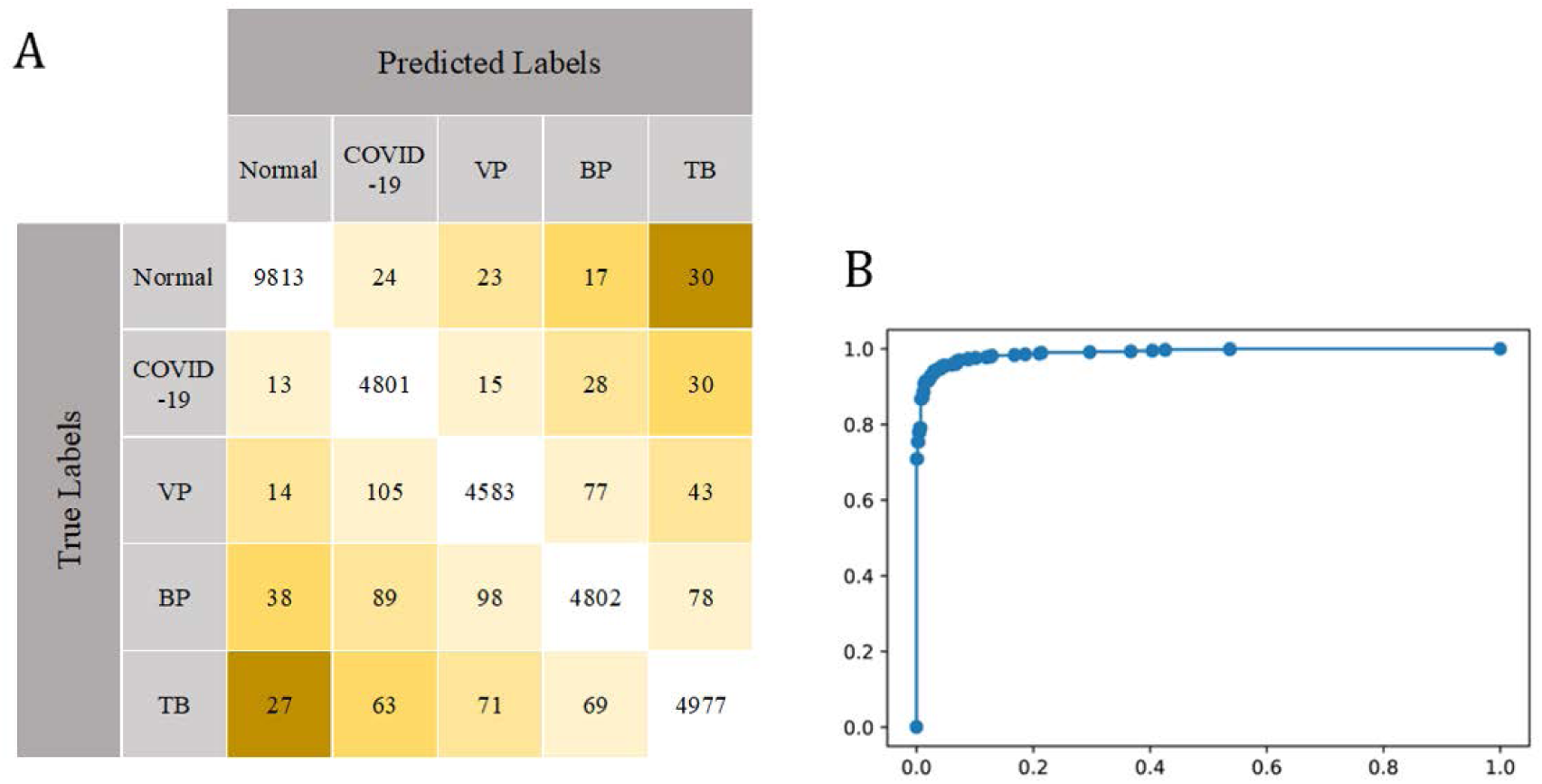
Performance of the AI algorithm during test. (A) Confusion matrix of the AI framework during test process. (B) ROC curve for binary classification between COVID-19 and the other four types of diseases or conditions.

## Discussion

Nowadays, COVID-19 has become global threaten. Timely diagnosis and isolation of including infected patients and asymptomatic carriers are critical to prevent further spread of the virus. RT-PCR-based detection of Coronavirus specific nucleic acid is regarded as the standard method of establishing the diagnosis of COVID-19. However, the positive rate of nucleic acid detection based on the samples collected from upper respiratory tract is unsatisfying. In an investigation with large sample, in the group of 0∼7 days after onset (d.a.o), positive rates based on the samples from throat swabs for severe cases and mild cases were 60.0% and 61.3%, respectively whereas positive rates for throat swabs reduced to 50.0% and 29.6% for severe cases and mild cases, respectively^(8)^.

COVID-19 may cause asymptomatic infection in some individuals^(9)^. Asymptomatic or mild cases combined are reported to represent about 40–50% of all infections^(10)^. Like confirmed COVID-19 cases, asymptomatic carriers of novel coronavirus acts as the infectious sources of COVID-19. Usually, confirmed COVID-19 patients are known risk and easy to prevent. However, asymptomatic carriers are “hidden enemies”, which tends to become mobile infectious sources.

Due to the unsatisfying positive rate nucleic acid detection and huge number of asymptomatic carriers, developing alternative methods of detection is urgently needed. Indeed, there is some significant advantages of detecting infected patients by CT scanning. According to the report by Ai *et al*,^(11)^ the positive rate of chest CT imaging is 88% for the diagnosis of suspected patients with COVID-19, which is superior to that of RT-PCR assay (59%).

Because of the relatively high positive rate of CT imaging in the early stage and the characteristic lesions of COVID-19 such as ground-glass opacity^(3, 4, 12)^, CT imaging has potential in the diagnosis of COVID-19 that cannot be ignored. There is large number of potential patients in need. More than that, each examination of CT imaging will generate a large number of images and significant inter-observer-variation exists in the interpretation of CT images. Thus, it is necessary to develop new auxiliary measures for the interpretation of CT images. In this study, we report an artificial intelligence framework based on deep learning for identifying COVID-19, which has balanced sensitivity and specificity. More than that, the area under the ROC curve was high as 99.0% evaluated by test dataset. Another advantage of our study is that five diseases or conditions were enrolled, which cover the most common infectious diseases of the lung. The limitation is that our AI framework will need further evaluation by more wide clinical application.

## Data Availability

None available

## Acknowledgements

This work was supported by the National Natural Science Foundation of China No.81871645 (J.S.P.). The funding source did not have any role in the design and conduct of the study; collection, management, analysis, and interpretation of the data; preparation, review, or approval of the manuscript; and decision to submit the manuscript for publication.

## Contributions

MF, and JSP conceived and designed the project. JSP obtained funding. QZ, SLY, YZ, YF, YL, XD, and YDR performed clinical diagnosis and collected samples. MF and JSP analysed and interpreted data. MF and LL trained and tested the AI model. JSP drafted the manuscript. JSP, QZ, and LL revised the manuscript. All the authors approved the final version of the manuscript.

